# Serum Brain-Derived Neurotrophic Factor Across the Human Lifespan: A Systematic Review and Meta-Analysis

**DOI:** 10.64898/2026.09.13.26362873

**Authors:** Zi Xin Wang, Xiuming Li, Hayley Dingsdale

## Abstract

**Background:** Bone marrow megakaryocytes, the primary source of brain-derived neurotrophic factor (BDNF) in the blood, secrete BDNF-packed platelets into the bloodstream. Reduced BDNF serum levels, a proxy for circulating blood-BDNF, have been linked to age-related neurological disorders, including dementia. Whether circulating BDNF levels decline with healthy aging or only in pathological contexts is unclear.

**Methods:** We searched PubMed, Embase, Web of Science, and ScienceDirect up to September 2025 for human studies reporting associations between age and serum BDNF in non-clinical populations. Fifteen studies were included. Effect sizes were extracted or converted as Pearson’s r and synthesized using a three-level random-effects meta-analysis, with subgroup analyses by age, heterogeneity, and quality.

**Findings:** Serum BDNF showed an age-specific decline. Among older adults (≥60 years), age was negatively associated with serum BDNF (pooled Fisher’s z=–0.139, 95% CI –0.166 to –0.111; p < 0.0001; I²=0.16%). In younger adults (<60 years), the association was not significant (pooled z=–0.098, 95% CI –0.344 to 0.148; p=0.435; I² = 87.2%). Across all ages, serum BDNF was inversely associated with age (pooled z=–0.127, 95% CI –0.245 to –0.009; p = 0.035), though heterogeneity was substantial (I² = 64.4%; Q (16) = 87.25, p < 0.0001).

**Interpretation:** Though not a feature of early or mid-adulthood, declining serum BDNF becomes pronounced in older age. This age-specific trajectory provides a reference point for future studies examining BDNF’s relation to cognitive aging and neurodegenerative risk.

**Funding:** No funding was received.

**Registration:** Open Science Framework - osf.io/264cp

## 1. Introduction

Brain-derived neurotrophic factor (BDNF) is one of the most extensively studied neurotrophic factors in biomedical research. Although most commonly associated with the brain, BDNF is primarily produced in the periphery, synthesized by megakaryocytes in the bone marrow and packaged into platelets, which release BDNF during coagulation and platelet activation (Chacón-Fernández et al., 2016). Emerging evidence from prospective and cross-sectional studies suggests that higher levels of blood-BDNF may exert a protective effect against progression to mild cognitive impairment (MCI) in cognitively healthy older adults (Kim et al., 2025) and lower serum BDNF has likewise been independently associated with age-related memory impairment in community-dwelling older adults (Mizoguchi et al., 2020).

Since ageing is associated with structural and functional decline in the brain (Peters, 2006; Schulz et al., 2022), circulating blood-BDNF has been investigated as a potential peripheral biomarker of cognitive health in older adults (Pisani et al., 2023). Several studies suggest that reduced BDNF levels in serum, a product derived from clotted blood, may correlate with age-related cognitive decline, hippocampal atrophy, and increased vulnerability to neurodegenerative conditions such as Alzheimer’s disease (Barde, 2025; Diniz & Teixeira, 2011; Erickson et al., 2010; Miranda et al., 2019). Lower serum BDNF has also been correlated with poorer memory performance and greater risk of cognitive impairment (Shimada et al., 2014), and has also been linked to cognitive deficits in metabolic conditions such as type 2 diabetes (Bathina & Das, 2015). However, baseline age-related patterns of peripheral BDNF remain poorly characterized (Naegelin et al., 2018).

To date, no systematic review has exclusively focused on the association between chronological age and serum BDNF concentrations across the human lifespan. Findings on the relationship between age and serum BDNF remain mixed; individual studies reported varying effect sizes and inconsistent directions of association, with some reporting negative correlations, others null effects, and some even positive associations (Kronenberg et al., 2021; Lang et al., 2004; Naegelin et al., 2018; Tunagur et al., 2024). This inconsistency is fueled by substantial variation in sample demographics, particularly the age range of the cohorts studied, as well as differences in measurement techniques and methodological approaches – making it difficult to establish a clear trajectory of serum BDNF across the lifespan.

To address this inconsistency and better define how BDNF levels change with age, we conducted the first systematic review and meta-analysis of studies reporting the association between chronological age and serum BDNF concentrations in humans. The objectives were to: (1) quantify the overall strength and direction of this relationship; (2) evaluate the extent of between-study heterogeneity; and (3) systematically investigate the sources of this heterogeneity using a pre-specified subgroup analysis based on cohort age. More specifically, this study seeks to provide a precise estimate of the age-BDNF association in healthy individuals and establish a robust effect size that can guide the design and interpretation of future research.

## 2. Methods

### 2.1. Reporting Standards

This systematic review and meta-analysis were guided by the Preferred Reporting Items for Systematic Reviews and Meta-Analyses (PRISMA) 2020 statement (see Supplementary Material for the completed PRISMA checklist). The project was initially conducted as a student research project for a national science fair, where preregistration of protocols is not typically required. For the purposes of subsequent journal submission and transparency, the protocol was registered retrospectively on the Open Science Framework (OSF; Registration URL: osf.io/264cp). The literature search, eligibility screening, and quality control were performed independently by two reviewers (ZW, XL) to minimize selection bias, with any discrepancies resolved through consensus-based discussion with a third reviewer (HD).

### 2.2. Search Strategy

To identify eligible human studies reporting the association between age and serum BDNF levels, we searched PubMed (MEDLINE), Elsevier (Embase), Web of Science, and ScienceDirect from inception to September 2025. Google Scholar was used only for backward/forward citation chasing and gray-literature discovery; it was not treated as a primary database due to reproducibility concerns which include lack of transparency and replicability, poor search precision and control, and potential bias towards popular English-language content. Full, line-by-line strategies and last-search dates for each database appear in Supplementary Table S1, along with per-database yields used in the PRISMA 2020 flow diagram.

### 2.3. Eligibility Criteria

#### 2.3.1 Inclusion

(1) **Population.** Human participants drawn from non-clinical/“healthy” samples. If mixed samples were reported (e.g., patient and control groups), only the healthy control subgroup was eligible. “Healthy” included community-dwelling individuals without diagnosed neurological or psychiatric disorders per study authors’ definitions. (2) **Exposure.** Chronological age treated as a continuous variable (or convertible to continuous). (3) **Outcome.** Serum BDNF measured with a validated immunoassay (e.g., ELISA, bead-based immunoassay). Particularly, studies measuring plasma or CSF BDNF were excluded. (4) **Effect Size.** Reported (or convertible) zero-order association between age and serum BDNF. Acceptable data included: Pearson’s r; statistics convertible to r from simple (age-only) linear regression (e.g., standardized β, t with df); or sufficient summary data to hand-calculate r. When necessary, if data were unavailable after two attempts at author contact, the study was excluded. (5) **Study design.** Cross-sectional studies; baseline (pre-intervention) data from cohorts or trials; or observational studies reporting the relevant association. (6) **Report characteristics.** Full-text, English-language, peer-reviewed articles published up to September 2025.

#### 2.3.2 Exclusion

(1) No extractable serum BDNF data, sample size N, effect sizes. (2) Alternative BDNF measures: plasma/CSF/whole blood. (3) Clinical samples or analyses without a healthy subgroup. (4) Duplicate data (overlapping cohorts – in which the report with the most complete data is retained) and unclear or inconsistent assay methodology precluding comparability. (5) Review Articles. (6) Reportings in languages other than English. (7) Animal studies.

We were strongly guided by the PRISMA 2020 guidelines for systematic reviews (Page et al., 2021). This study selection process is summarized in the PRISMA flowchart (Figure 1). All study exclusions and inclusions were done manually, with no use of artificial intelligence throughout the entire selection process.

**Figure 1.**
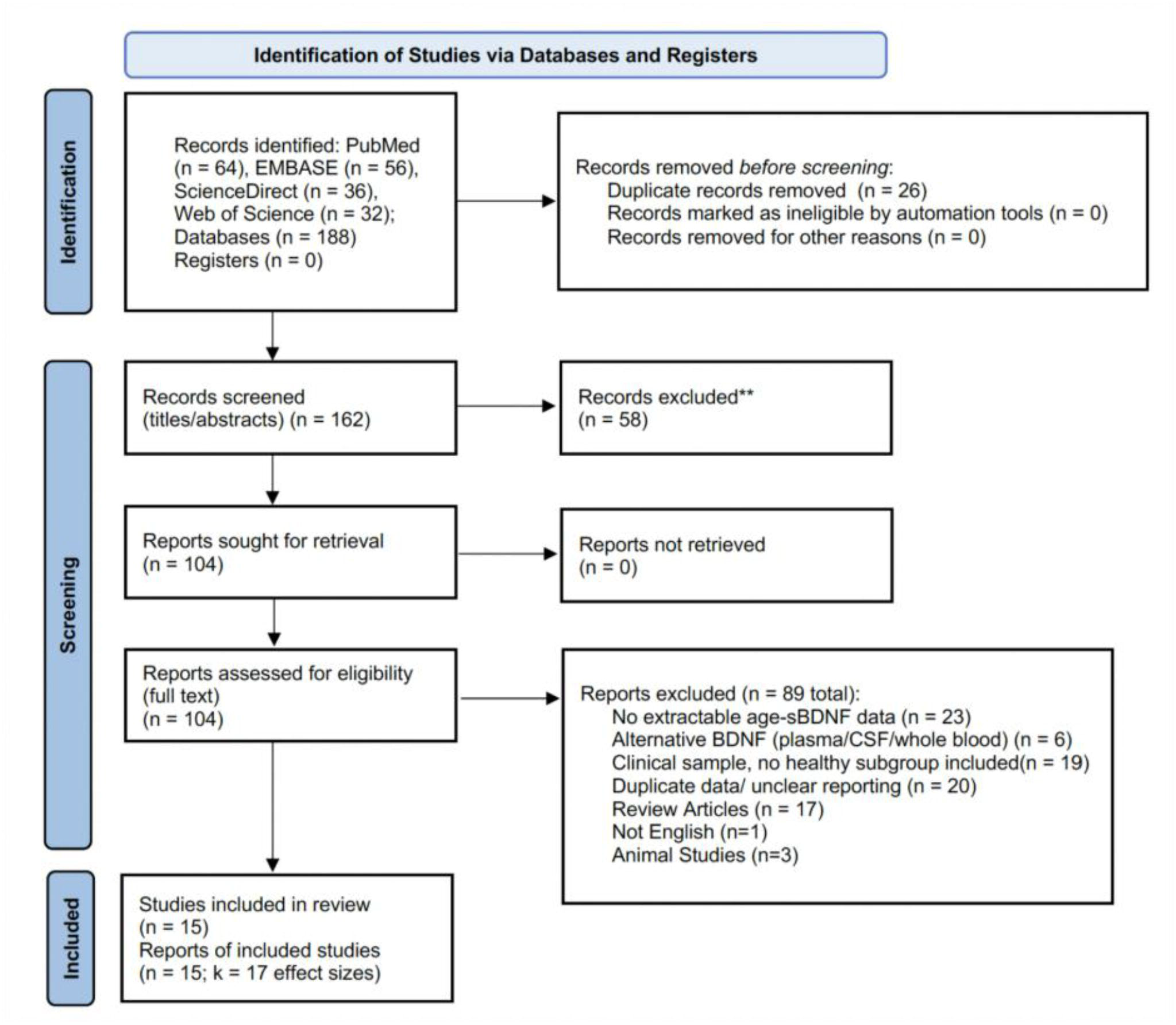
PRISMA 2020 flow diagram of study selection.

### 2.4. Quality Assessment

All three authors evaluated the methodological quality of included studies using the Joanna Briggs Institute (JBI) Critical Appraisal Checklist for Analytical Cross-Sectional Studies (Joanna Briggs Institute., 2020). Criteria assessed included sample representativeness, measurement validity, completeness of reporting, and control for confounders. See Supplemental Material (Table S2).

### 2.5. Data Extraction

For each eligible study, we extracted: **(a)** Correlation coefficient (Pearson’s r) between age and serum BDNF **(b)** Sample size (N) **(c)** Mean age **(d)** Percentage of female participants, and **(e)** Continent for statistical analysis. We extracted bivariate Pearson’s r when only non-parametric correlations were reported. When only single-predictor standardized β or t(df) from simple linear regression was available, we converted it to r using standard formula. We did not derive r from multi-predictor models. When studies reported both proBDNF and mature BDNF (mBDNF), we extracted mBDNF only because it is the biologically active isoform and best matches the results quantified in the rest of the literature; proBDNF was excluded beforehand.

### 2.6. Statistical Analysis

All analyses were conducted in R version 4.5.1 (R Core Team 2025) using the metafor package (version 4.8). (Viechtbauer, 2010). Effect sizes were extracted as Pearson’s correlation coefficients (r).

To account for the statistical dependence of multiple effect sizes derived from the same study (e.g., sex-stratified subgroups), we used a three-level random-effects meta-analysis model for the primary analysis, with effect sizes nested within studies (Cheung, 2014).

Between-study heterogeneity was quantified using the Q-statistic and the I² statistic. To investigate the sources of heterogeneity, we performed a pre-specified subgroup analysis based on mean cohort age (’Younger Cohort’: <60 years; ’Older Cohort’: ≥60 years). We also conducted univariable meta-regression to test the percentage of female participants and continents as potential moderators. All statistical tests were two-tailed, with p < 0.05 considered significant.

We dichotomized cohorts at 60 years for three reasons. Firstly, 60 is a widely used epidemiologic threshold for “older adult” status in ageing research and aligns with how several of our included studies reported demographics (Shimada et al., 2014; World Health Organization, 2025; UNHCR, 2025). Second, by 60 years nearly all women are postmenopausal, reducing large, hormone-related variability in circulating BDNF that characterizes the perimenopausal decade (Pluchino et al., 2009); this improves interpretability of age-group contrasts. Third, with platelet count and platelet function changing with age, and platelets being major contributors to measured serum BDNF, a >= 60 stratification has a biological plausibility (Balduini & Noris, 2014; Chacón-Fernández et al., 2016; Le Blanc & Lordkipanidzé, 2019).

It is important to note that the study with no reported mean age (Shimada et al., 2014) had a demographic that guaranteed its classification in the ‘Older Cohort’ group, with the participants’ age range being 65-97.

## 3. Results

### 3.1. Study Characteristics

Our search identified 188 records, of which 15 studies (providing 17 effect sizes) met our inclusion criteria (Fig. 1). Sample sizes across these effect sizes ranged from 14 to over 2000. The meta-analysis included a total of 5797 healthy individuals. Study characteristics, including sample size, demographics, and BDNF measurement method, are detailed in Table 1. Overall, the mean JBI score was 7.4/8, with a median 8/8 (IQR 7-8); 9/15 (60%) were rated Low risk of bias, with no studies rated high. See Supplemental Material (Table S2).

**Table 1:**
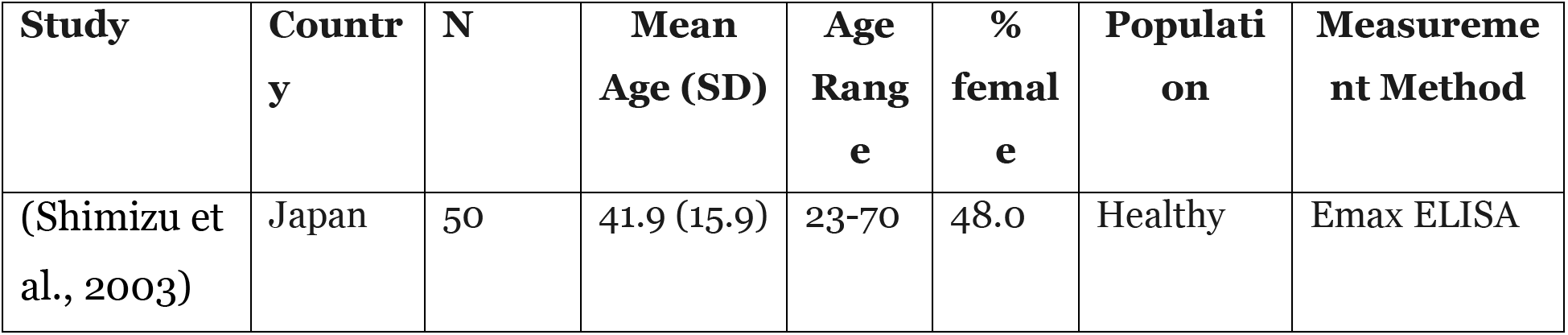
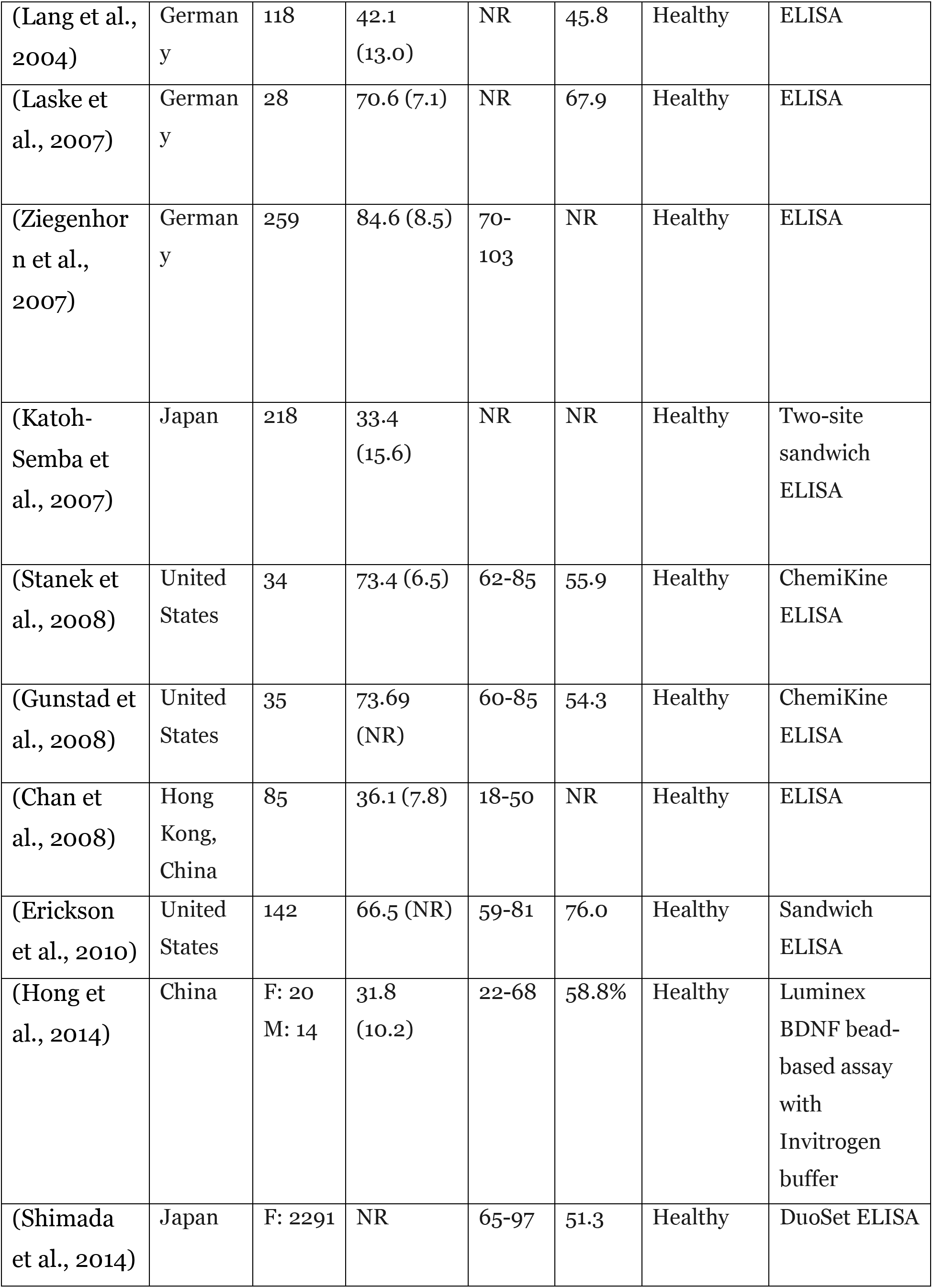
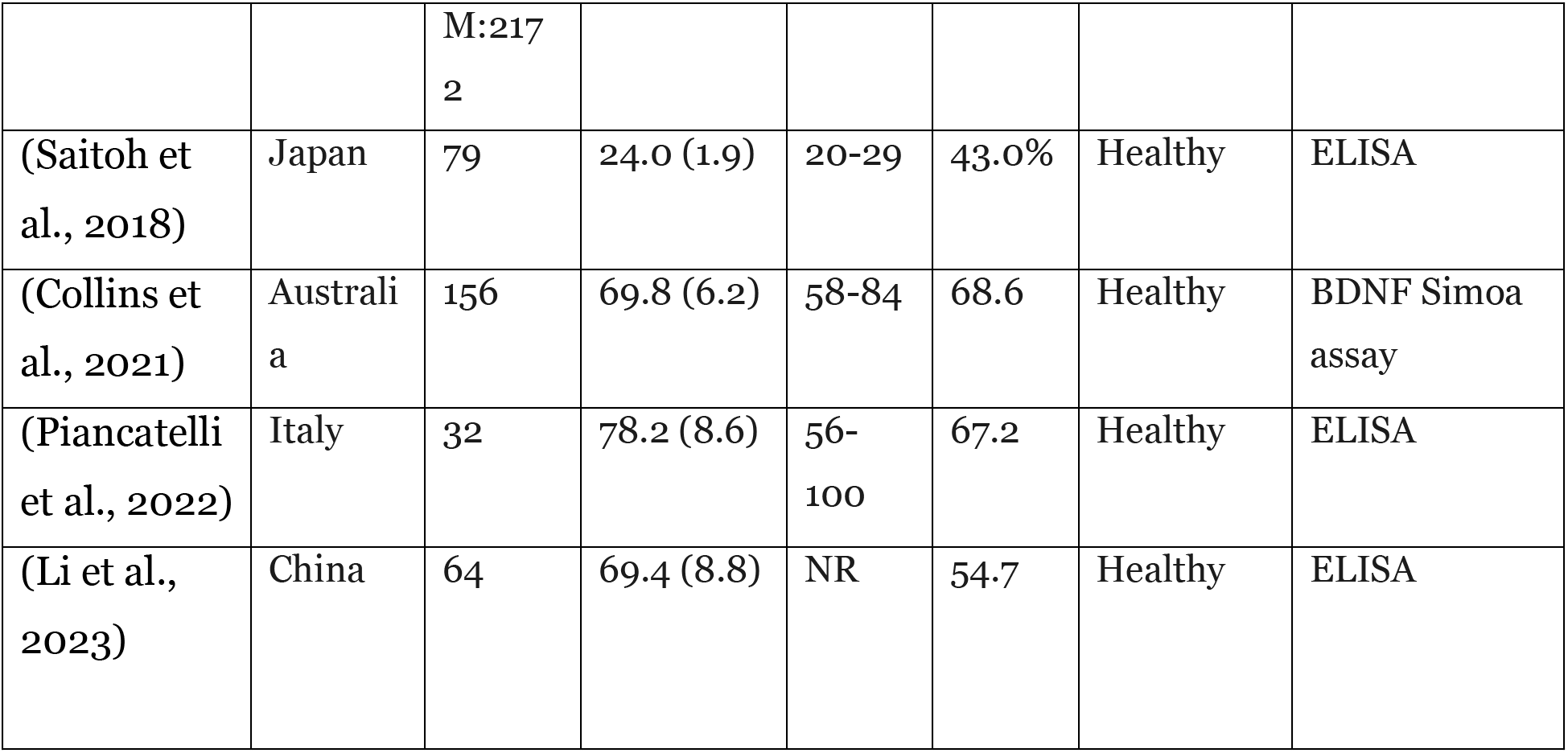
Characteristics of included studies (sorted by year of publication). Where only sex-stratified results were available (Hong et al., 2014^a^ and Shimada et al., 2014^b^), subgroup sizes are shown for each row and M/F contribute to two dependent effect sizes. Dependence among multiple effects from the same study was modeled using a three-level random-effects framework with variance components at the study and effect levels; effects were not treated as independent in the pooled analysis. For Li et al., 2023^c^, only mBDNF (mature/biologically active BDNF) was extracted and analyzed; proBDNF was not used, and mBDNF was treated as the serum BDNF outcome. “Measurement method” denotes the assay/platform used to quantify serum BDNF. NR = not reported; ELISA = enzyme-linked immunosorbent assay; mBDNF = mature BDNF.

### 3.2. Primary Meta-analysis Reveals a Modest Overall Effect with High Heterogeneity

Using a three-level random-effects model, the pooled Fisher’s Z was -0.127 (95% CI: [-0.245, -0.009], p = 0.035), corresponding to a pooled Pearson’s correlation coefficient of r = -0.126. This indicates a modest but statistically significant inverse relationship between age and serum BDNF levels (Figure 2). However, this overall effect was characterized by substantial and significant between-study heterogeneity (I² = 64.4%; Q (df=16) = 87.25, p < 0.0001).

**Figure 2:**
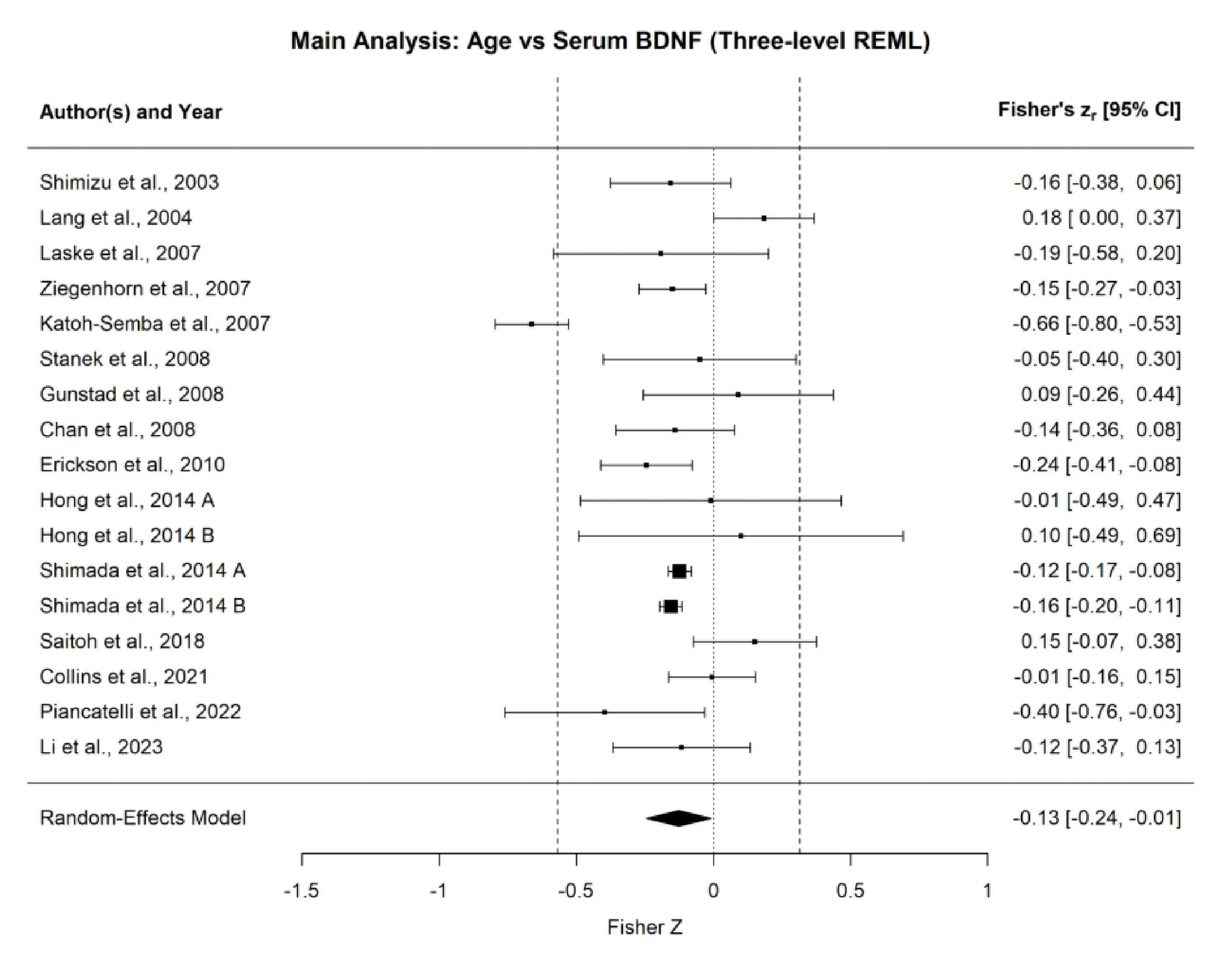
Forest plot illustrating individual study effect sizes (Fisher’s z) and the overall pooled estimate from the three-level random-effects model. Each square represents the effect size of an individual study, with its size proportional to the study’s weight (inverse-variance) in the analysis. The diamond represents the overall pooled estimate. The pooled estimate indicates a modest but statistically significant inverse association between age and serum BDNF (pooled z = -0.127, p = 0.035).

The prediction interval was –0.514 to 0.305 on the r scale (back-transformed). Furthermore, analyses for publication bias and leave-one-out sensitivity testing confirmed the robustness of the overall dataset, with no evidence of small-study effects or undue influence from any single study (see Supplementary Material, Figures S1-S3).

### 3.3. Subgroup Analysis by Cohort Age

To investigate the source of this heterogeneity, we conducted a subgroup analysis based on the mean age of the study cohorts (Figure 3).

**Figure 3:**
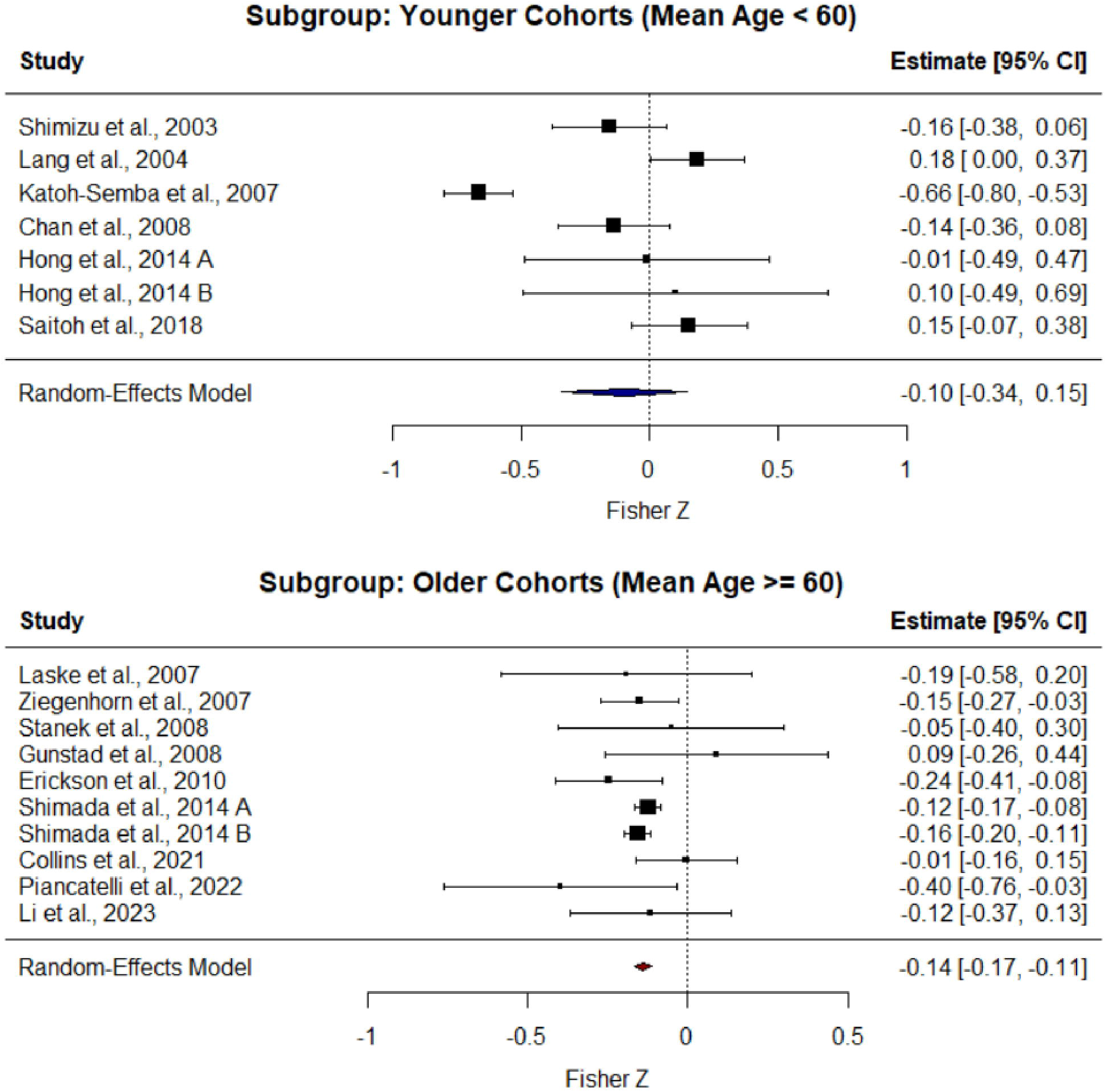
Subgroup analysis of the association between age and serum BDNF. (Top) In studies with a mean cohort age below 60 years, the pooled effect size was not significant, and heterogeneity remained high. (Bottom) In studies with a mean cohort age of 60 years or older, the pooled effect size was highly significant and negative, with heterogeneity eliminated.

In the ’Younger Cohort’ subgroup (k=7 effects; mean age < 60), there was no significant overall association between age and serum BDNF (pooled Z = -0.098, 95% CI [-0.344, 0.148], p = 0.43). Heterogeneity within this subgroup remained extremely high (I² = 87.16%).

In stark contrast, the ’Older Cohort’ subgroup (k=10 effects; mean age ≥ 60) revealed a consistent and highly significant negative association between age and serum BDNF (pooled Z = -0.139, 95% CI [-0.166, -0.111], p < 0.0001). Crucially, heterogeneity within this older group was effectively eliminated (I² = 0.16%). These results demonstrate that the overall heterogeneity is driven by inconsistent findings in younger cohorts, and that the age-related decline in BDNF is a robust phenomenon specific to later life.

### 3.4 Moderators

Beyond age composition, the examined moderators did not explain additional heterogeneity. Continent/region was non-significant (QM (df = 2) = 0.226, p = 0.893; reference = Asia; levels tested: Asia, Europe, North America), as was assay platform (QM (df = 1) = 0.382, p = 0.537; reference = ‘ELISA’, vs ‘Luminex’). The percentage of female participants (per 10-percentage-point increase) was also non-significant (QM (1) = 0.220, p = 0.639; β = −0.007, 95% CI −0.037 to 0.023). These findings support the interpretation that cohort age structure, rather than sampling region, assay type, or sex balance, is the dominant driver of heterogeneity.

## 4. Discussion

### 4.1. A Life-Stage Dependent Relationship

This study is the first meta-analysis to systematically examine the relationship between chronological age and serum brain-derived neurotrophic factor (BDNF) concentrations. By synthesizing 17 effect sizes derived from 15 studies, this work provides a quantitative summary of a research question that, until now, has only been examined through individual studies. Employing a random-effects model using restricted maximum likelihood (REML) estimation, this meta-analysis revealed a statistically significant negative correlation between age and serum BDNF concentrations (Fisher’s z = –0.127, 95% CI –0.245 to –0.009; p = 0.035). Our findings provide strong evidence that may help reconcile previously conflicting reports in the literature. In younger cohorts (<60 years), the absence of a consistent association and the presence of high heterogeneity suggest that serum BDNF levels during this life stage are shaped by a range of dynamic factors that can obscure a subtle ageing effect. In younger cohorts, age-dependent hormonal fluctuations (e.g., estrogen variation in females before menopause) (Scharfman & MacLusky, 2006) represent a unique source of variability. In addition, factors such as acute and chronic stress (Barde, 2025) and differences in lifestyle, including diet and physical activity, can influence circulating BDNF across the lifespan. By contrast, the striking consistency and near-complete elimination of heterogeneity observed in older cohorts (≥60 years) suggest that, beyond this threshold, the cumulative effects of biological ageing emerge as the dominant regulator of serum BDNF. This shift allows the underlying trajectory of age-related decline to be observed with greater clarity and reliability. None of the tested moderators (continent, assay platform, or sex balance) accounted for the between-study variability, reinforcing that age composition is the primary determinant of the observed heterogeneity.

This finding remained consistent and robust across multiple analytical procedures. The leave-one-out sensitivity testing confirmed that no single study disproportionately influenced the pooled effect. Additionally, all bias tests suggested reduced risk of small-study bias as well as the consistency in the direction of the observed association.

However, while this analysis identifies a statistically significant negative correlation between age and serum BDNF, it is important to note that it does not establish a causal relationship. The observed overall decline might instead be due to unmeasured factors, such as alcohol consumption (Shafiee et al., 2023), blood glucose levels (Davarpanah et al., 2021) and the effect of the environment across lifespan (Kobiec et al., 2023).

Nonetheless, this work fills in a critical gap in the current field by providing the first quantitative synthesis of all available evidence on the relationship between serum BDNF and age.

### 4.2. Context within the Literature

The overall conclusion that serum BDNF levels decline with chronological age is supported by many studies in the field. For example, platelet counts are known to decrease with age, a finding highlighted by (Balduini & Noris, 2014), who suggest this reduction may be a key factor underlying the observed negative association between age and serum BDNF. However, some studies have reported a positive correlation between serum BDNF and age. These contrasting findings suggest that factors beyond platelet number may contribute to serum BDNF levels. In particular, Zahavi et al. (Zahavi et al., 1980) reported that platelet reactivity and the release of α-granule components such as PF4 increase with age, which may help explain positive correlations between serum BDNF and age despite reduced platelet counts.

### 4.3. Clinical Implications and Biological Context

This observation is of potential clinical relevance as previous research has linked lower serum BDNF concentrations with cognitive decline and an increased risk of mild cognitive impairments (MCI). A large cross-sectional study of older adults reported that individuals with serum BDNF levels 1.5 standard deviations below the mean exhibited poorer memory performance and were more likely to meet the MCI criteria (Shimada et al., 2014). However, more research, especially longitudinal studies, must be done to further confirm these findings. It has also been suggested that decreased serum BDNF levels may have a role in the pathophysiology of the cognitive deficits observed in patients with type 2 diabetes mellitus (Bathina & Das, 2015). Additional research has also shown a possible negative correlation between the presence of Parkinson’s disease and concentrations of BDNF in human serum (Scalzo et al., 2010). Conversely, individuals with higher serum BDNF levels have also been shown to have a lower risk of developing dementia (Weinstein et al., 2014), suggesting a possible correlation between concentrations of serum BDNF and the disease. Although these studies suggest possible associations, the biological context makes clinical interpretation more difficult, as serum BDNF is derived mainly from platelets, which reflect megakaryocytic activity in the bone marrow (Barde, 2025). Because of this, these peripheral measures are shaped by hematological and metabolic factors rather than mirroring neural tissue physiology (Barde, 2025). To date, no clear evidence supports the diffusion of BDNF across the blood-brain barrier (Barde, 2025). Serum BDNF has nonetheless been implicated in several neurological and psychiatric conditions, including mild cognitive impairment (MCI), depression, and potentially Alzheimer’s disease (Kim et al., 2025; Ng et al., 2019). However, findings in these areas remain inconsistent, and much more research is required to resolve these disputes. Due to this, serum BDNF levels, while accessible, cannot be proven to accurately reflect neurotrophin concentrations within the human brain. For these observed relationships between chronological age and serum BDNF to have greater clinical relevance, further research is needed to directly investigate BDNF expression in central nervous system tissues. Such studies will be essential to clarify the relevance of peripheral BDNF concentrations in terms of brain ageing and neurodegeneration.

### 4.4. Sources of Heterogeneity and Limitations

The primary strength of this study is that subgrouping by mean cohort age explains the bulk of the observed heterogeneity: overall I² = 64.4% collapses to 0.16% within the ≥60 year stratum, while heterogeneity remains high (I² = 87.16%) in the <60 year stratum.

Thus, age composition, rather than publication bias, appears to be the dominant driver of inconsistency in the literature.

Several additional limitations should also be noted. The first being the reliance on cross-sectional data; statistically, this design captures only a single snapshot in time and thus reveals individual correlations between age and serum BDNF rather than the longitudinal trajectory of BDNF decline. Genetic variation also remains an underexplored source of heterogeneity. The most widely studied of these is Val66Met (rs6265), which impairs activity dependent BDNF release by 18-30% in the brain compared to Val/Val homozygotes (Portaccio, 2021). This allele is common: approximately 30-50% of Caucasian individuals carry (Pivac et al., 2009), and up to 70% of Asian individuals (China, Japan, Korea) carry at least one Met allele (Bian et al., 2005). Met/Met homozygotes are relatively uncommon in Caucasians (1-4%) but occur at higher frequencies in Asians (∼20%) (Shen et al., 2018). Genotype-specific effects add further variability and inconsistencies in terms of BDNF levels in the periphery. The effects of Val/Met heterozygotes have been mixed, but evidence suggests that Met/Met homozygotes often have lower reported serum BDNF levels (Ozan et al., 2010), though the pattern is not uniform across all populations. The high prevalence of this polymorphism and its impact on BDNF levels in the central nervous system and periphery underscores the need to account for genetic polymorphisms in future studies. Doing so would likely reduce heterogeneity, allowing a sharper interpretation of age-related trends in serum BDNF (Brown et al., 2020). Other lesser-known polymorphisms such as cleavage site polymorphisms rs1048220 and rs1048222 may also exert important effects (Barde, 2025).

Finally, as reflected in the JBI table, there remains some ambiguity in the definition of “healthy” control samples, as inclusion criteria varied across studies and were not consistently standardised. This inconsistency may have further contributed to the heterogeneity and reduced comparability of values across studies.

### 4.5. Future Work

Although this meta-analysis provides robust evidence for an age-related decline in serum BDNF, the high degree of heterogeneity highlights a significant need for methodological refinement in future research. A priority should be standardizing BDNF measurement protocols, particularly regarding pre-analytical variables such as clotting time and centrifugation strategy, including centrifugal force and time, for both serum and plasma extraction methods. Variability in these factors has been shown to significantly impact BDNF quantification, as highlighted by (Gejl et al., 2019), who reported that both processing protocols and storage time had effects on altering measured BDNF levels in peripheral blood samples from both serum and plasma.

Consistent methodologies across laboratories, particularly for ELISA preparation and sample handling, are critical for ensuring data comparability and reducing noise in meta-analytical outcomes.

As of now, the correlation between chronological age and serum BDNF has indeterminate clinical implications for neurological health, due to the understanding that peripheral BDNF levels do not reliably reflect central nervous system concentrations (Barde, 2025). Therefore, to make age-related declines in serum BDNF relevant as a biomarker for neurological health, it is necessary to explore alternative BDNF measurement sources that have stronger and more consistent associations with central values. One promising avenue currently being explored is salivary BDNF, where recent findings indicate a possible link between measured concentrations and BDNF released by sensorimotor neurons of the peripheral nervous system (Barde, 2025; Mandel et al., 2009). This line of research may represent an important step toward developing methods capable of approximating brain BDNF levels, which have been shown in prior studies to serve as correlates of neurological health, especially concerning synaptic function and neuroplasticity.

## 5. Concluding Remarks

This meta-analysis found a modest but statistically significant negative association between chronological age and serum BDNF levels across 15 studies (n = 5,797 participants, age range 18-103 years). The decline in serum BDNF was primarily observed in older adults (≥60 years), where the relationship was consistent and homogeneous, while studies involving younger adults (<60 years) showed no clear pattern and substantial heterogeneity. These findings indicate that measurable age-related decreases in circulating BDNF emerge mainly in later adulthood.

Although the overall correlation is small, this work provides a quantitative reference for understanding serum BDNF variation across the human lifespan. Future research should focus on standardizing assay protocols and determining whether peripheral BDNF accurately reflects central neurotrophic activity related to cognitive ageing.

## Data Availability

All data produced in the present study are available upon reasonable request to the authors.

## Acknowledgements

The authors gratefully acknowledge Professor Yves-Alain Barde for his informal review and insightful comments. We also thank a research associate from the University of British Columbia for their mentorship and detailed feedback on the preliminary statistical analyses.

## Author Contributions (CRediT)

ZW and XL are recognized as co-first authors. All authors approved the final version of this manuscript.

Conceptualization was carried out by ZW. Methodology and investigation were jointly conducted by ZW and XL. Data curation was completed by XL and ZW. Visualization, including the PRISMA flow diagram and schematic layout, was performed by ZW. Formal analysis involving statistical modelling and meta-analysis was conducted by XL and ZW, also with guidance from a research associate from UBC. Validation, including sensitivity, bias, and robustness checks, was performed by XL under guidance from a research associate from UBC. Visualization of the meta-analytic figures, including forest and funnel plots, was completed by XL with additional guidance. The initial draft of the manuscript was written by ZW and XL, and project administration was jointly handled by both authors. HD contributed to manuscript review and provided supervision. No external resources, funding, or software were used.

