## Supplementary Materials for "Serum Brain-Derived Neurotrophic Factor Across the Human Lifespan: A Systematic Review and Meta-Analysis"

**This PDF file includes:**

Methodology
- Search Clauses
- MeSH terms for PubMed

Raw Data Collected

Study Characteristics for Selected Studies

JBI RoB
- Detailed justification of RoB

### Methodology

Search clauses

We searched PubMed (MEDLINE), Embase (Elsevier), ScienceDirect, and Web of Science from inception through September 2025. Google Scholar was used for backward/forward citation chasing and grey-literature discovery only. Full, reproducible strategies (including field tags and controlled vocabulary), per-database yields, and last-search dates are provided in Table S1.

**Table S1.** Electronic database search (most recent search date: 2nd Sept 2025). The methodological quality and risk of bias for each included study were independently assessed by two reviewers (ZW, XL) using the Joanna Briggs Institute (JBI) Critical Appraisal Checklist for Analytical Cross-Sectional Studies. A third reviewer (HD) resolved any discrepancies to reach a consensus. The checklist evaluates studies based on eight key criteria to determine the internal validity and risk of bias. Studies were scored based on the number of criteria they met, with a higher score indicating a lower risk of bias.

| **INFORMATION SOURCE** | **PUBMED (MEDLINE)** |
|---|---|
| **SEARCH STRATEGY** | (("Aging"[MeSH] OR "Age Factors"[ MeSH] OR "age-related"[tiab] OR "chronological age"[tiab] OR lifespan[tiab])<br>AND ("Brain-Derived Neurotrophic Factor"[Mesh] OR BDNF[tiab])<br>AND (serum[tiab] OR "Blood"[ MeSH] OR "circulating BDNF"[tiab]))<br>NOT (animal[MeSH] NOT human[MeSH]) |
| **NO. CONSIDERED FOR META** | 64 |
| **LAST SEARCH** | Sept 2nd 2025 |

| **INFORMATION SOURCE** | **EMBASE (ELSEIVER)** |
|---|---|
| **SEARCH STRATEGY** | #1 'brain derived neurotrophic factor'/exp OR bdnf:ti,ab,kw OR 'brain-derived neurotrophic factor':ti,ab,kw<br><br>#2 'blood serum'/exp OR serum:ti,ab,kw OR |

| | |
|---|---|
| | ( (bdnf OR 'brain-derived neurotrophic factor') NEAR/3 serum ):ti,ab,kw<br>/* captures “serum BDNF” */<br><br>#3 #1 AND #2<br><br>#4 'age'/exp OR 'aged'/exp OR 'aging'/exp OR 'chronological age'/exp OR 'life span'/exp OR<br>(age:ti,ab,kw OR aged:ti,ab,kw OR aging:ti,ab,kw OR<br>'chronological age':ti,ab,kw OR (age NEAR/1 related):ti,ab,kw OR lifespan:ti,ab,kw)<br><br>#5 'cognition'/exp OR 'biological marker'/exp OR 'dementia'/exp OR<br>(cognit*:ti,ab,kw OR biomarker*:ti,ab,kw OR dementia*:ti,ab,kw)<br><br>#6 #3 AND #4 AND #5<br><br>#7 #6 NOT ('animal'/exp NOT 'human'/exp) /* exclude animal-only records */<br><br>#8 #7 AND [english]/lim<br>#9 #7 AND [article]/lim |
| **NO. CONSIDERED FOR META** | 56 |
| **LAST SEARCH** | Sept 2$^{nd}$ 2025 |

| **INFORMATION SOURCE** | **SCIENCEDIRECT** |
|---|---|
| **SEARCH STRATEGY** | "age" AND "serum BDNF" |
| **NO. EXAMINED FULL TEXT** | 36 |
| **LAST SEARCH** | Sept 1$^{st}$ 2025 |

| **INFORMATION SOURCE** | **WEB OF SCIENCE** |
|---|---|
| **SEARCH STRATEGY** | "TS=("serum BDNF" OR ("serum" AND "brain-derived neurotrophic factor"))<br>AND<br>TS=(age OR aging OR "chronological age" OR lifespan OR elderly OR adolescent OR youth OR child*) |

| | |
|---|---|
| | NOT<br>TS=(animal* OR rat OR mouse OR rodent OR "in vitro") |
| **NO. CONSIDERED FOR META** | 32 |
| **LAST SEARCH** | Sept 2nd 2025 |

MeSH terms for PubMed

Age Factors

Aged

Aging / blood*

Biomarkers/ blood

Brain / physiopathology

Brain-Derived Neurotrophic Factor / blood*

Brain-Derived Neurotrophic

### Raw Data Collected

Study Characteristics for Selected Studies

| Study | Country | r | N | Mean Age (SD) | Age Range | % female | Population | Measurement Method |
|---|---|---|---|---|---|---|---|---|
| *Shimizu et al., 2003* | Japan | -0.155 | 50 | 41.9 (15.9) | 23-70 | 48.0 | Healthy | Emax ELISA |
| *Lang et al., 2004* | Germany | 0.182 | 118 | 42.1 (13.0) | NR | 45.8 | Healthy | ELISA |
| *Laske et al., 2007* | Germany | -0.189 | 28 | 70.6 (7.1) | NR | 67.9 | Healthy | ELISA |
| *Ziegenhorn et al., 2007* | Germany | -0.149 | 259 | 84.6 (8.5) | 70-103 | NR | Healthy | ELISA |
| *Katoh-Semba et al., 2007* | Japan | -0.58 | 218 | 33.4 (15.6) | NR | NR | Healthy | Two-site sandwich ELISA |
| *Stanek et al., 2008* | United States | -0.05 | 34 | 73.4 (6.5) | 62-85 | 55.9 | Healthy | ChemiKine ELISA |
| *Gunstad et al. 2008* | United States | 0.09 | 35 | 73.69 (NR) | 60-85 | 54.3 | Healthy | ChemiKine ELISA |
| *Chan et al., 2008* | Hong Kong, China | -0.139 | 85 | 36.1 (7.8) | 18-50 | NR | Healthy | ELISA |
| *Erickson et al., 2010* | United States | -0.24 | 142 | 66.5 (NR) | 59-81 | 76.0 | Healthy | Sandwich ELISA |
| *Hong et al., 2014*[a] | China | -0.01<br>0.1 | F: 20<br>M: 14 | 31.8 (10.2) | 22-68 | 58.8% | Healthy | Luminex BDNF bead-based assay with Invitrogen buffer |
| *Shimada et al., 2014*[b] | Japan | -0.123<br>-0.154 | F: 2291<br>M:2172 | NR | 65-97 | 51.3 | Healthy | DuoSet ELISA |
| *Saitoh et al., 2018* | Japan | 0.15 | 79 | 24.0 (1.9) | 20-29 | 43.0% | Healthy | ELISA |
| *Collins et al., 2021* | Australia | -0.005 | 156 | 69.8 (6.2) | 58-84 | 68.6 | Healthy | BDNF Simoa assay |
| *Piancatelli et al., 2022* | Italy | -0.377 | 32 | 78.2 (8.6) | 56-100 | 67.2 | Healthy | ELISA |
| *Li et al., 2023*[c] | China | -0.116 | 64 | 69.4 (8.8) | NR | 54.7 | Healthy | ELISA. |

<u>Small-study bias and sensitivity analyses (with a three-level primary model)</u>

Rationale. Our primary meta-analysis uses a three-level random-effects model (REML) to address statistical dependence among multiple effects within a study (e.g., sex strata). On the other hand, common small-study and influence diagnostics – funnel plots, Egger's regression, trim-and-fill, and leave-one-out (LOO) - are defined for single-level models that assume independence. Following standard practice, we therefore report these diagnostics from single-level random-effects sensitivity models, while keeping the three-level results as our primary inference.

Implementation overview.

1. Funnel/Egger/Trim-and-fill (single-level RE): run on the same effect sizes used in the three-level analysis (Fisher's z), treating them in a standard single-level model for diagnostic purposes.
2. Study-cluster LOO (single-level RE): to respect within-study dependence during LOO, we delete all effects from one Study_ID at a time, re-fit a single-level REML model on the remaining data, and record the pooled estimate after each deletion. That is, if a study has two effect sizes due to sex stratification, both will be deleted.

*3.1 Funnel Plot & Egger's regression (single-level sensitivity)*

We fit a single-level random-effects model to the Fisher's z effects and produced a funnel plot (z vs SE). Egger's test for small-study effects was non-significant ($z = 0.83$, $p = 0.41$).

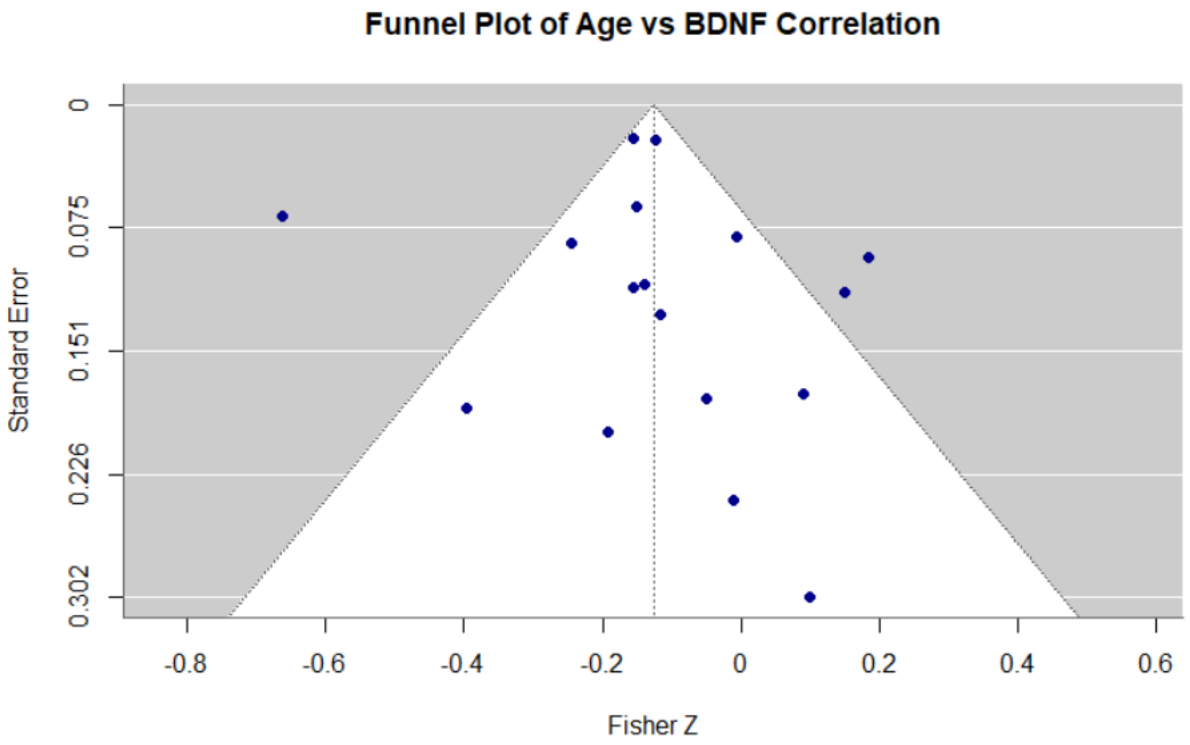


**Figure S1:** Funnel of Fisher's z versus standard error from the single-level random-effects sensitivity model. Egger's test: $z = 0.83$, $p = 0.41$ (no evidence of small-study effects).

*3.2 Trim-and-fill (single-level sensitivity)*

Trim-and-fill (left-side) imputed 6 studies, yielding an adjusted pooled Fisher's $z = -0.220$ (SE 0.055; 95% CI –0.328 to –0.111). After imputation the model heterogeneity was $Q(22) = 145.19$, $I^2 = 91.39\%$, $\tau^2 = 0.0515$. This adjustment shifts the magnitude but does not change the direction of the association.

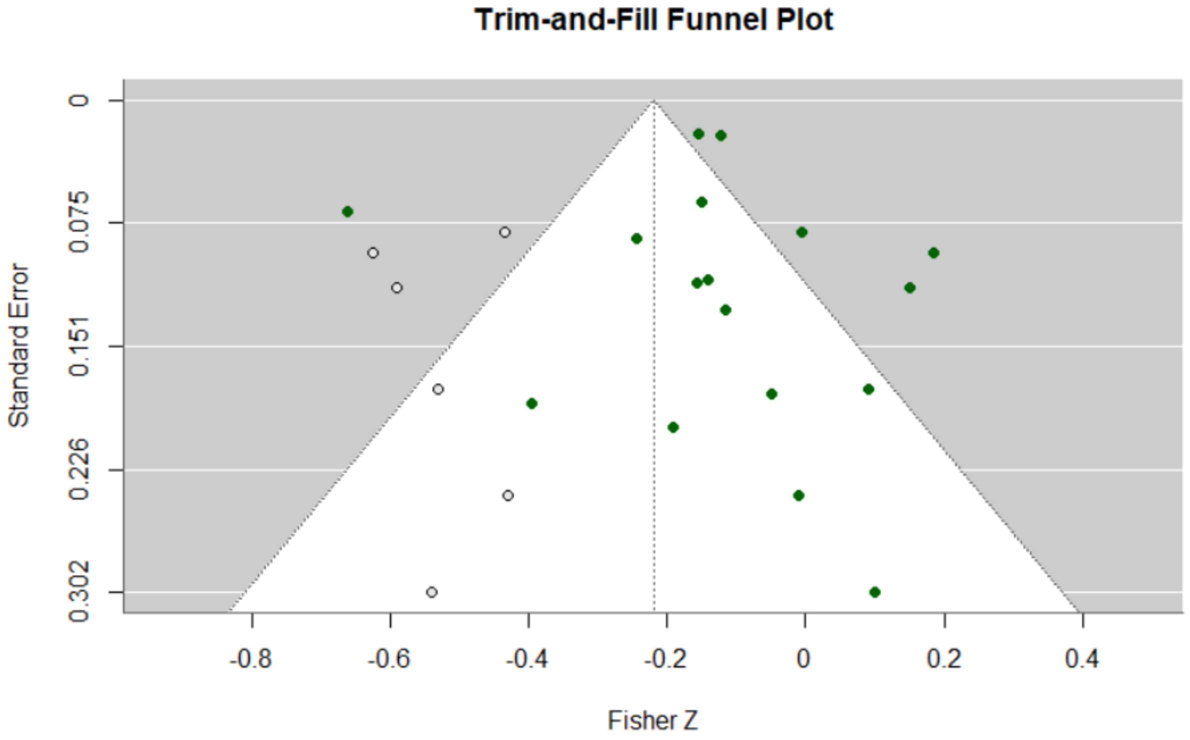


**Figure S2:** Trim-and-fill imputed 6 studies (left), adjusted pooled Fisher's z = –0.220 (95% CI –0.328 to –0.111). Heterogeneity after imputation: $Q(22)=145.19$, $I^2=91.39\%$, $\tau^2=0.0515$.

*3.3 Study-cluster leave-one-out (single-level REML)*

To ensure deletions respect within-study dependence, we performed study-cluster LOO, removing all effects belonging to one Study_ID at a time and re-fitting a single-level REML model to the remaining data. Across deletions, the back-transformed pooled correlation (r) ranged from –0.149 to –0.091, indicating that no single study drove the overall negative association.

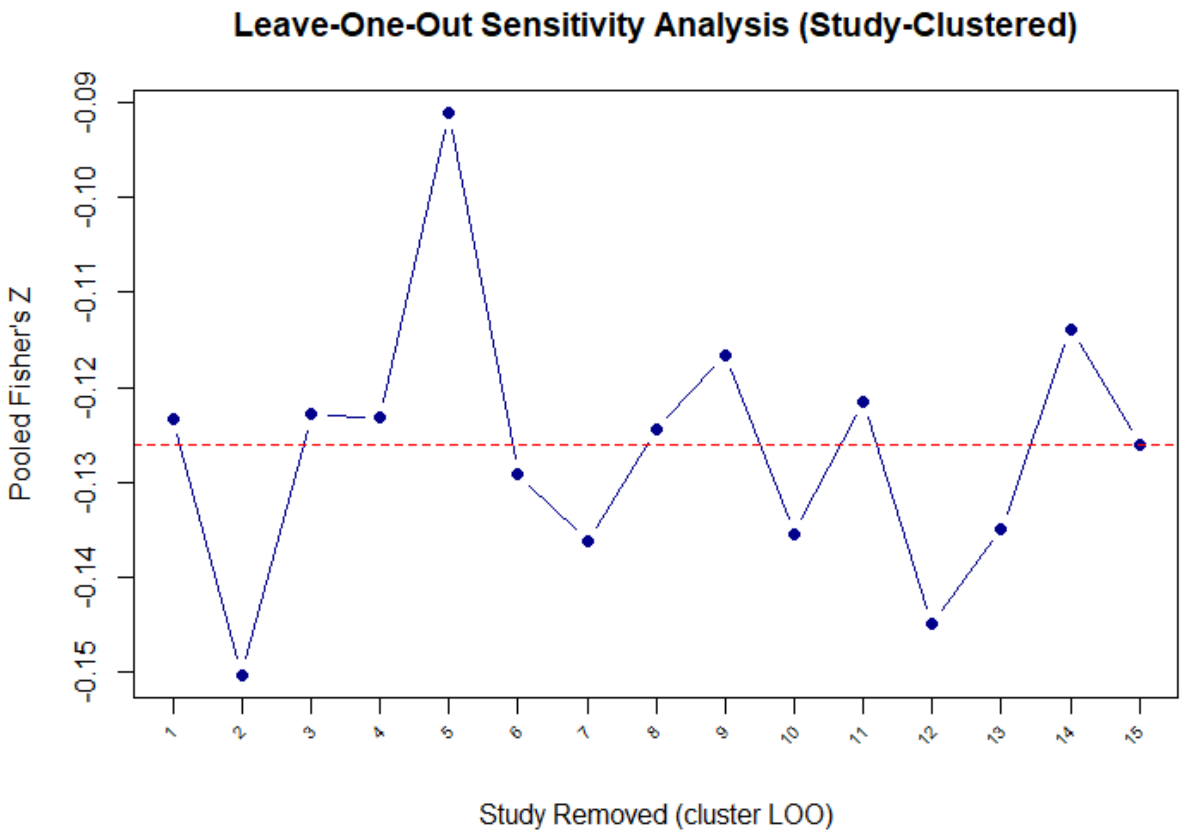


**Figure S3:** Each point shows the pooled Fisher's z after removing all effects from one study. The back-transformed pooled r ranged from –0.149 to –0.091 across deletions. The dashed horizontal line denotes the pooled estimate from the all-studies single-level sensitivity model.

*3.4 Reporting Note*

The three-level REML model remains the primary analysis reported in the Main text (overall Fisher's z, 95% CI, $I^2$, Q, prediction interval, and pre-specified subgroups). The single-level diagnostics above serve as complementary checks for small-study effects and study influence.

### Reviewer 1 Joanna Briggs Institute Risk of Bias Assessment (Methodological Quality Assessment)

**Table S2.** Risk of Bias Summary Table 1

| AUTHORS | Q1 | Q2 | Q3 | Q4 | Q5 | Q6 | Q7 | Q8 | TOTAL | RISK LEVEL |
|---|---|---|---|---|---|---|---|---|---|---|
| **SHIMIZU ET AL., 2003** | ✓ | ✓ | ✓ | ✓ | ✓ | -- | ✓ | ✓ | 7/8 | Low |
| **LANG ET AL., 2004** | ✓ | ✓ | ✓ | ✓ | ✓ | ✓ | ✓ | ✓ | 8/8 | Low |
| **LASKE ET AL., 2007** | ✓ | ✓ | ✓ | ✓ | ✓ | -- | ✓ | ✓ | 7/8 | Low |
| **ZIEGENHORN ET AL., 2007** | ✓ | ✓ | ✓ | ✓ | ✓ | -- | ✓ | ✓ | 7/8 | Low |
| **KATOH-SEMBA ET AL., 2007** | ✓ | ✓ | ✓ | ✓ | ✓ | -- | ✓ | ✓ | 7/8 | Low |
| **STANEK ET AL., 2008** | ✓ | ✓ | -- | ✓ | ✓ | -- | ✓ | ✓ | 6/8 | Moderate |
| **GUNSTAD ET AL. 2008** | ✓ | ✓ | ✓ | --- | ✓ | ✓ | ✓ | ✓ | 7/8 | Low |
| **CHAN ET AL., 2008** | ✓ | --- | --- | --- | --- | ✓/- | ✓ | ✓ | 3.5/8 | High |
| **ERICKSON ET AL., 2010** | ✓ | ✓ | ✓ | ✓ | --- | --- | ✓ | ✓ | 6/8 | Moderate |
| **HONG ET AL., 2014** | ✓ | ✓ | ✓ | ✓ | ✓ | ✓ | ✓ | ✓ | 8/8 | Low |

| | | | | | | | | | | |
|---|---|---|---|---|---|---|---|---|---|---|
| **SHIMADA ET AL., 2014** | ✓ | ✓ | ✓ | ✓ | ✓ | ✓ | ✓ | ✓ | 8/8 | Low |
| **SAITOH ET AL., 2018** | ✓ | ✓ | ✓ | ✓ | ✓ | ✓ | ✓ | ✓ | 8/8 | Low |
| **COLLINS ET AL., 2021** | ✓ | ✓ | ✓ | ✓ | --- | --- | ✓ | ✓ | 8/8 | Low |
| **PIANCATELLI ET AL., 2022** | ✓ | ✓ | ✓ | --- | ✓ | --- | ✓ | ✓ | 6/8 | Moderate |
| **LI ET AL., 2023** | ✓ | ✓ | ✓ | ✓ | --- | --- | ✓ | ✓ | 6/8 | Moderate |

Q.1) Were the criteria for inclusion in the sample clearly defined?

Q.2) Were the study subjects and the setting described in detail?

Q.3) Was the exposure measured in a valid and reliable way?

Q.4) Were objective, standard criteria used for measurement of the condition?

Q.5) Were confounding factors identified?

Q.6) Were strategies to deal with confounding factors stated?

Q.7) Were the outcomes measured in a valid and reliable way?

Q.8) Was appropriate statistical analysis used?

Abbreviations: Yes: ✓, No/Unclear: ---

Low: 100-80%, Moderate: 80-50%, High 50-20%

https://www.researchgate.net/figure/JBI-critical-appraisal-quality-score-and-summary-of-conclusions-of-the-retrieved-reviews_tbl1_264708110

**Reviewer 2 XL:**
**Detailed Justification of Risk of Bias (Methodological Quality Assessment)**

**Table S3.** Risk of Bias Summary Table 2

| AUTHORS | Q1 | Q2 | Q3 | Q4 | Q5 | Q6 | Q7 | Q8 | TOTAL | RISK LEVEL |
|---|---|---|---|---|---|---|---|---|---|---|
| **SHIMIZU ET AL., 2003** | ✓ | ✓ | ✓ | ✓ | ✓ | ✓ | ✓ | ✓ | 8/8 | Low |
| **LANG ET AL., 2004** | ✓ | ✓ | ✓ | ✓ | ✓ | ✓ | ✓ | ✓ | 8/8 | Low |
| **LASKE ET AL., 2007** | ✓ | ✓ | ✓ | ✓ | ✓ | ✓ | ✓ | ✓ | 8/8 | Low |
| **ZIEGENHORN ET AL., 2007** | ✓ | ✓ | ✓ | ✓ | ✓ | ✓ | ✓ | ✓ | 8/8 | Low |
| **KATOH-SEMBA ET AL., 2007** | ✓ | ✓ | --- | --- | ✓ | ✓ | ✓ | ✓ | 6/8 | Moderate |
| **STANEK ET AL., 2008** | ✓ | ✓ | ✓ | --- | ✓ | ✓ | ✓ | ✓ | 7/8 | Low-Moderate |
| **GUNSTAD ET AL. 2008** | ✓ | ✓ | ✓ | --- | ✓ | ✓ | ✓ | ✓ | 7/8 | Low-Moderate |
| **CHAN ET AL., 2008** | ✓ | ✓ | ✓ | --- | ✓ | ✓ | ✓ | ✓ | 7/8 | Low-Moderate |
| **ERICKSON ET AL., 2010** | ✓ | ✓ | ✓ | ✓ | ✓ | ✓ | ✓ | ✓ | 8/8 | Low |
| **HONG ET AL., 2014** | ✓ | ✓ | ✓ | ✓ | ✓ | ✓ | ✓ | ✓ | 8/8 | Low |

| | | | | | | | | | | |
|---|---|---|---|---|---|---|---|---|---|---|
| **SHIMADA ET AL., 2014** | ✓ | ✓ | ✓ | ✓ | ✓ | ✓ | ✓ | ✓ | 8/8 | Low |
| **SAITOH ET AL., 2018** | ✓ | ✓ | ✓ | ✓ | ✓ | ✓ | ✓ | ✓ | 8/8 | Low |
| **COLLINS ET AL., 2021** | ✓ | ✓ | ✓ | ✓ | ✓ | ✓ | ✓ | ✓ | 8/8 | Low |
| **PIANCATELLI ET AL., 2022** | --- | ✓ | ✓ | --- | ✓ | ✓ | ✓ | ✓ | 6/8 | Moderate |
| **LI ET AL., 2023** | --- | ✓ | ✓ | --- | ✓ | ✓ | ✓ | ✓ | 6/8 | Moderate |

Q.1) Were the criteria for inclusion in the sample clearly defined?

Q.2) Were the study subjects and the setting described in detail?

Q.3) Was the exposure measured in a valid and reliable way?

Q.4) Were objective, standard criteria used for measurement of the condition?

Q.5) Were confounding factors identified?

Q.6) Were strategies to deal with confounding factors stated?

Q.7) Were the outcomes measured in a valid and reliable way?

Q.8) Was appropriate statistical analysis used?

Abbreviations: Yes: ✓, No/Unclear: ---

Low: 100-80%, Moderate: 80-50%, High 50-20%

https://www.researchgate.net/figure/JBI-critical-appraisal-quality-score-and-summary-of-conclusions-of-the-retrieved-reviews_tbl1_264708110
